# A Statistical Analysis on COVID-19 Pandemic in the City of Toronto

**DOI:** 10.1101/2022.12.28.22284001

**Authors:** Ali R. Kaazempur-Mofrad

## Abstract

As with many viruses, a main topic of concern that has arisen during the COVID-19 pandemic is the nature of the virus’ spread and the impact it presents for varying age groups. Using statistical methods and numerical summaries, I analysed the impact that age has on the incubation period, death rates, and case outcomes of COVID-19 cases in the city of Toronto. The results of this study illustrate that the elderly have shorter incubation periods, higher death rates, and a higher count of cases which result in fatality. The analysis concludes that the impact of COVID-19 is most felt by the elderly.

## 1. Introduction

With the COVID-19 pandemic reaching the front pages of just about every corner of the world, evaluating its impact in the city of Toronto can help get a better understanding on the virus’s general behavior and interaction within a large community. While every part of the world has felt the impact of the pandemic on different levels, Toronto is the chosen city for this case study.

This statistical analysis focuses on exploring the impacts of COVID-19 on various age groups. In this analysis, the primary objectives are to investigate how age impacts the incubation period, death rate, and case outcomes.

## 2. Data

In order to conduct this study, data was obtained from the city of Toronto’s `opendatatoronto` library [1]. This database is extracted from the provincial Case & Contact Management System (CCM) and is published by Toronto Public Health on a weekly basis. Given that this database is still being updated every Wednesday, the data reflected in this analysis is accurate as of *July 15, 2021*.

### 2.1. Data Cleaning

After collecting the data from the library and prior to evaluating the data, a cleaning process was conducted, as follows.

First, in order optimize the data and make it compatible for use in R, the variable names needed to be converted to remove spaces and punctuation. Next, because some observations within the database didn’t have complete information and had been left blank (NA), those observations were ignored in order to prevent errors in the analysis process. Lastly, to ensure the database was entirely unique, a quick test for duplicate data was conducted which yielded no duplicate entries.

### 2.2. Key Variables

In their dataset publication, Toronto Public Health took into consideration several different factors including demographic, geographic, and severity information for each case.

While the database features several different variables such as `source_of_infection` and `neighbourhood_name`, this study focuses primarily on the `episode_date`, `reported_date`, `age_group` and `outcome` variables.

- `episode_date` - the estimated date when the disease was acquired. Due to the uncertainty of the exact date of contamination, this variable lists the earliest date between when symptoms are expressed, test collection date, and the reported date. The date format is yyyy-mm-dd.
- `reported_date` - the date when each case was reported to Toronto Public Health. The date format is yyyy-mm-dd.
- `age_group` - age of contaminated individuals at the time of illness. This variable is *categorical* rather than *numerical* and has the following age group breakdown:

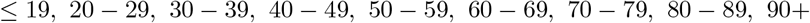
- `outcome` - current outcome of each case at the moment of the study. The 3 possible options for this variable are *active, fatal*, and *resolved*.
- `days_until_report` - reflects the difference, in days, between the dates from `episode_date` and `reported_date`
- `death` - a dummy variable (also referred to as a binary variable) that evaluates inputs from the `outcome` variable and sets values based on the following breakdown:

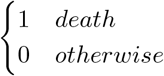

## 3. Methods

During the COVID-19 pandemic, being able to identify and efficiently report cases has proven vitally important. The period of time between the moment an individual contracts the COVID-19 virus (denoted hereafter as “*episode date*” according to Toronto Public Health [1]) and when the individual is informed and has their case reported (denoted hereafter as “*report date*” per Toronto Public Health) is a dangerous interval where an infected individual can unknowingly spread the virus onto others.

Knowing the interval between the episode date and the report date helps to get a better understanding of how long it takes for symptoms to show in infected individuals. Of course, the data could be skewed by individuals who delay reporting their symptoms for various reasons. However, for the case of this study, we will *assume* that the interval between the episode date and report date is an indication of how long it takes for symptoms to show.

## 4. Results

In this section, I present statistical models and data summaries to investigate the trends of COVID-19 incubation periods, death rates, and case outcomes all with respect to varying age groups.

### 4.1. Maximum Likelihood Estimation Analysis

In order to gain a better understanding of how quickly and efficiently new cases are reported (i.e. the interval between the episode date and report date), we will create a model to compare the rates of reporting cases across different age groups in Toronto.

To fit a model for the analysis of the estimated number of days between the episode date and report date, a *univariate frequentist* model can be used. The data collected by Toronto Public Health is used as sample data to form an estimation model. We will use an *exponential distribution* because the data that is being evaluated reflects the amount of time until a case is reported. The model estimation used for this purpose is:

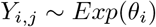

where *Y*_*i,j*_ is the average number of days for a case to be reported since infection for each age group. On the other hand, *θ*_*i*_ is the case reporting rate for each age group.

In the histogram below (*Figure 1*), the number of days between episode date and report date is displayed for all cases reported to Toronto Public Health.

**Figure 1:**
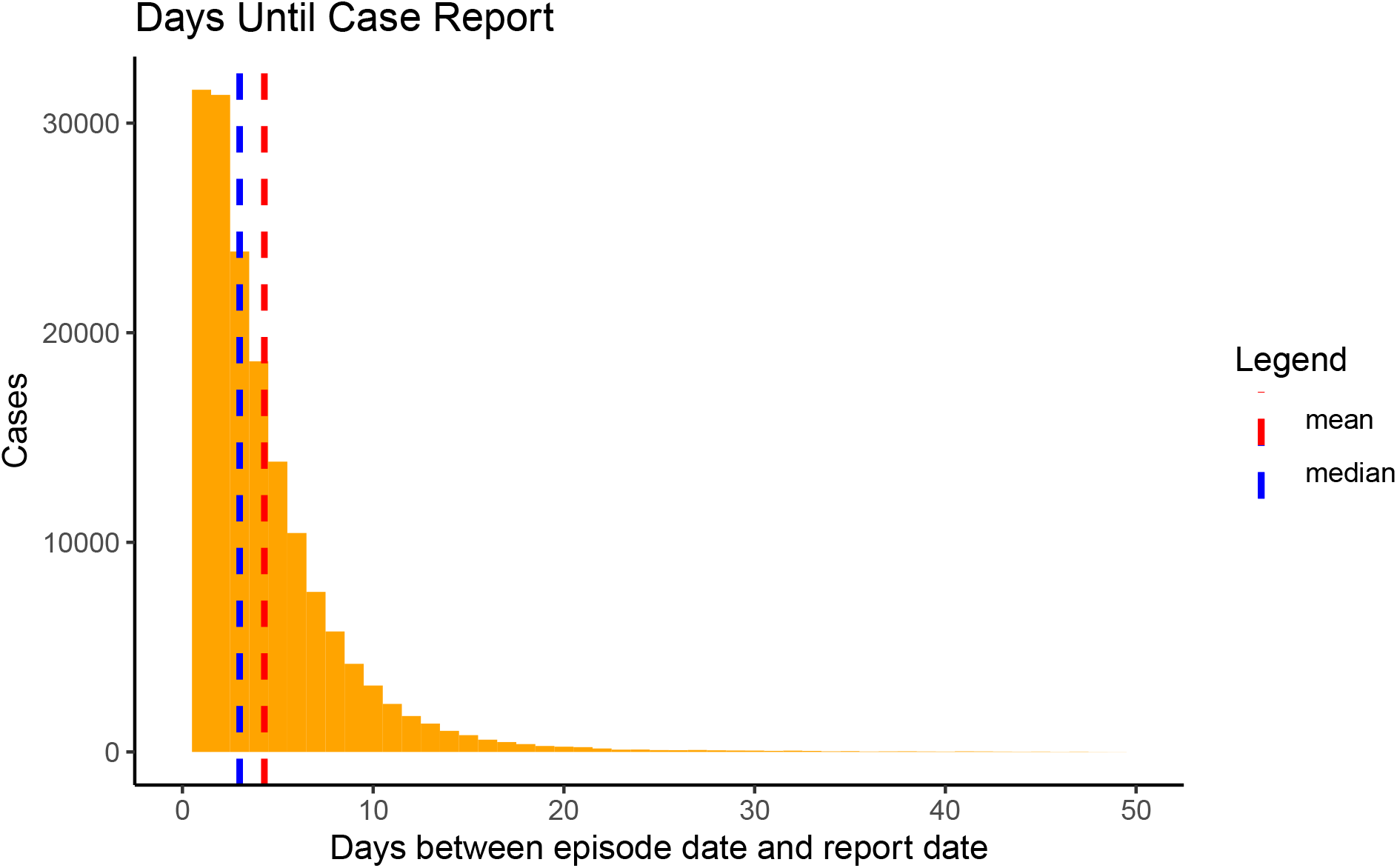
Number of days for a case to be reported following the episode date (defined as the date an individual contracts the COVID−19 virus). As evident in this histogram, the data decays exponentially as a large majority of cases are typically reported within the first few days.

With this model estimation, we can find the event rate of reporting a case per day.

| cases | deaths | death_rate |
| --- | --- | --- |
| 167389 | 3536 | 0.0211244 |

| Age Group | Report Rate (Case/Day) |
| --- | --- |
| 19 and younger | 0.2550655 |
| 20 to 29 Years | 0.2296172 |
| 30 to 39 Years | 0.2287716 |
| 40 to 49 Years | 0.2264382 |
| 50 to 59 Years | 0.2259619 |
| 60 to 69 Years | 0.2211251 |
| 70 to 79 Years | 0.2330952 |
| 80 to 89 Years | 0.2544146 |
| 90 and older | 0.2819221 |

In the table above, it is evident that the general population (across all ages) has a rate of reporting around 0.25 cases per day which equates to approximately an average of around 4 days since the episode date.

It appears that individuals 90 years and older have the highest rate of reporting a case per day. This suggests that for the elderly, the symptoms of COVID-19 likely appear sooner than younger individuals.

Aside from the elderly, it appears that youth at ages 19 and younger also have a higher rate of reporting cases per day. This may possibly be due to weaker immune systems of the children which results in a shorter period of time for symptoms to flare up.

Another noticeable aspect of the data is that individuals between the age groups of 20 to 29 and 60 to 69 years have lower rates of reporting cases per day. This might be explained by individuals within these age groups having a more developed immune system than children and the elderly. Another potential cause for this lower rate could be that individuals within these age groups are typically busier with work and are more prone to delay reporting of symptoms

As evident in Figure 1, it takes an *average (mean)* of **4.296 days** from the day which the disease is contracted until the day that the case is reported to Toronto Public Health. Because the mean is prone to outliers in the dataset, the *median* of **3 days** is likely a better measurement of the data.

The spread of the days it takes to report a case is described by a *variance* of **26.06** with a *standard deviation* of **5.11 days**.

### 4.2. Numerical Summaries: COVID-19 Death Rates

Using the new dummy variable of `death`, a separate dataset `covid_death_ages` was created to highlight the breakdown of the number of deaths, cases, and death rate for each age group reflected original dataset.

As per the most recent update to the dataset since July 15, 2021, there have been 167,389 COVID-19 cases fully filed in Toronto. Amongst these cases, there have been a total of 3,536 fatalities which amounts to a 2.11% fatality rate amongst all age groups.

In order to see whether the death rate varies across different ages, the data was broken down into several age groups. As per the table below, the fatality rate from COVID-19 increases with age as the rate is *lowest* in the *19 and younger* age group and *highest* in the *90 and older* age group.

To see this relationship visually, the scatterplot below (*Figure 2*) also shows that the fatality rate increases with each increase in age group.

| age_group | deaths | cases | death_rate |
| --- | --- | --- | --- |
| 19 and younger | 1 | 24459 | 0.0000409 |
| 20 to 29 Years | 10 | 34176 | 0.0002926 |
| 30 to 39 Years | 23 | 28763 | 0.0007996 |
| 40 to 49 Years | 54 | 24199 | 0.0022315 |
| 50 to 59 Years | 177 | 24142 | 0.0073316 |
| 60 to 69 Years | 417 | 15184 | 0.0274631 |
| 70 to 79 Years | 691 | 7570 | 0.0912814 |
| 80 to 89 Years | 1213 | 5763 | 0.2104807 |
| 90 and older | 950 | 3133 | 0.3032237 |

**Figure 2:**
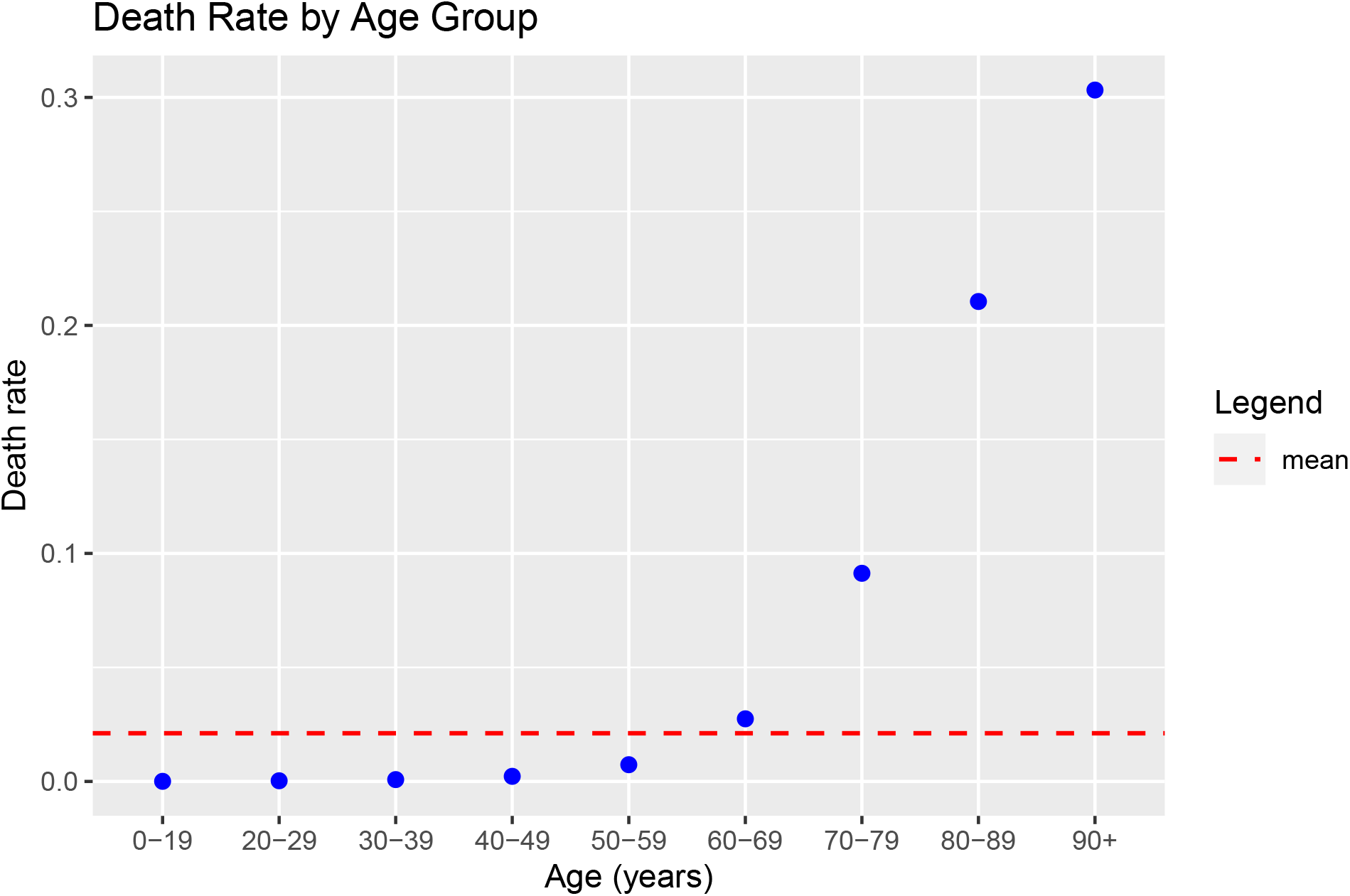
Death rate for each age group in Toronto

The scatterplot in Figure 2 also shows the fatality rate of 2.11% across all age groups (red dashed line), as determined earlier. This enables viewers to clearly see that the elders face a fatality rate well above the average rate while individuals in the 50-59 and 60-69 age groups are closest to the average.

### 4.3. Case Outcomes by Age Group

Rather than focusing entirely on the fatality rate, it is also important to analyze the total counts of each outcome in the dataset (*active, fatal*, and *resolved*). As highlighted in the bar graphs below (*Figure 3*), the majority of the fatalities in Toronto occurred in the elderly (as evident by the left skew in the plot). On the other hand, the amount of resolved cases in younger individuals is far greater than those in the elderly (as evident by the right skew in the plot).

**Figure 3:**
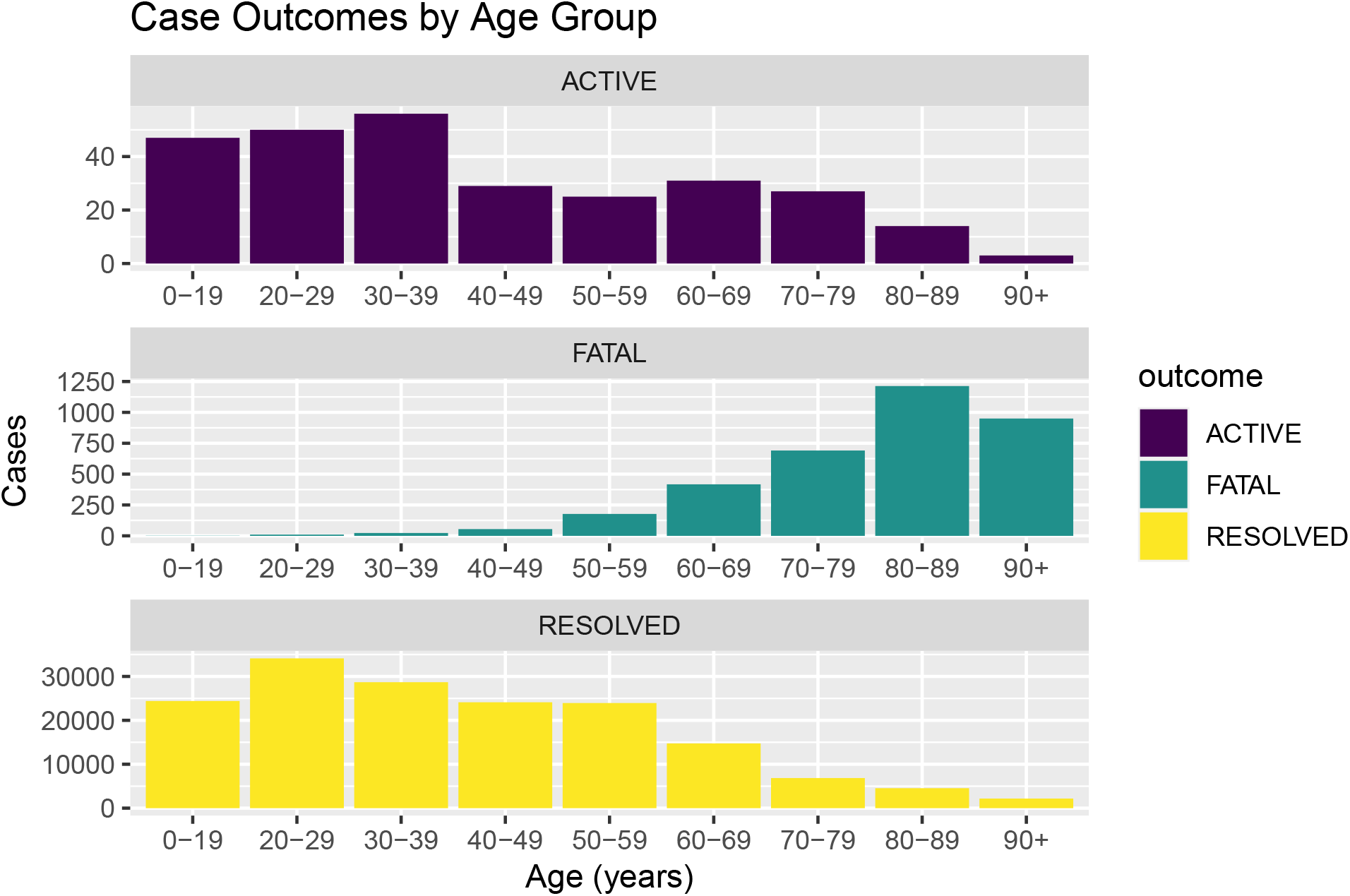
Instances of each case outcome by age group − Top: Active, Middle: Fatal, Bottom: Resolved

The figure also shows that the active cases are spread in more of a normal distribution which could imply that the virus is still spreading across all age groups, although the total amount of active cases is not as vast as earlier in the pandemic period.

## 5. Conclusion

This study investigated the impact age has on COVID-19 cases. To understand the relationship between age and incubation period, death rate, and case outcomes, I used statistical methods such as maximum likelihood estimations and numerical summaries.

The results of this study showed that the incubation period, or the period between the episode date and the report date, was shortest for the elderly and children under the age of 19. This could indicate that the weaker immune systems in children and the elderly result in COVID-19 symptoms appearing earlier than other age groups.

This study also led to understanding that the recovery rate of COVID-19 is significantly higher in younger individuals while the elderly are more prone to fatality.

One major limitation of this dataset is that we are not given any information regarding other health conditions that patients might have. Due to this, the study is limited to focus primarily on age without being able to investigate other factors which may impact case outcomes.

To further this investigation, it would be helpful to obtain further information on each COVID-19 case in Toronto. This would enable the study to tackle other potential causes and factors that could potentially impact the results of this analysis.

## Data Availability

All data produced are available online at:
https://open.toronto.ca/dataset/covid-19-cases-in-toronto/

